# Prediction of Overall Patient Characteristics that Incorporate Multiple Outcomes in Acute Stroke: Latent Class Analysis

**DOI:** 10.1101/2023.05.24.23290504

**Authors:** Junya Uchida, Moeka Yamada, Hirofumi Nagayama, Kounosuke Tomori, Kohei Ikeda, Keita Yamauchi

## Abstract

**Background:** Previous prediction models have predicted a single outcome (e.g. gait) from several patient characteristics at one point (e.g. on admission). However, in clinical practice, it is important to predict an overall patient characteristic by incorporating multiple outcomes. This study aimed to develop a prediction model of overall patient characteristics in acute stroke patients using latent class analysis.

**Methods:** This retrospective observational study analyzed stroke patients admitted to acute care hospitals (37 hospitals, N=10,270) between January 2005 and March 2016 from the Japan Association of Rehabilitation Database. Overall, 6,881 patients were classified into latent classes based on their outcomes. The prediction model was developed based on patient characteristics and functional ability at admission. We selected the following outcome variables at discharge for classification using latent class analysis: Functional Independence Measure (functional abilities and cognitive functions), subscales of the National Institutes of Health Stroke Scale (upper extremity function), length of hospital stay, and discharge destination. The predictor variables were age, Functional Independence Measure (functional abilities and comprehension), subscales of the National Institutes of Health Stroke Scale (upper extremity function), stroke type, and amount of rehabilitation (physical, occupational, and speech therapies) per day during hospitalization.

**Results:** Patients (N=6,881) were classified into nine classes based on latent class analysis regarding patient characteristics at discharge (class size: 4–29%). Class 1 was the mildest (shorter stay and highest possibility of home discharge), and Class 2 was the most severe (longer stay and the highest possibility of transfers including deaths). Different gradations characterized Classes 3–9; these patient characteristics were clinically acceptable. Predictor variables at admission that predicted class membership were significant (odds ratio: 0.0– 107.9, *P*<.001).

**Conclusions:** Based on these findings, the model developed in this study could predict an overall patient characteristic combining multiple outcomes, helping determine the appropriate rehabilitation intensity. In actual clinical practice, internal and external validation is required.

## Introduction

Annually, 15 million people worldwide suffer from stroke, resulting in five million deaths and five million developing stroke-related disabilities.^1)^ Rehabilitation services are included as the indispensable mechanism that encourages functional recovery and independence in acute stroke patients.^2)^ A formal assessment of activities of daily living (ADLs), instrumental ADLs, communication abilities, and functional mobility is recommended for the rehabilitation of stroke patients before discharge. The findings should be incorporated into the care transition and discharge planning process.^2, 3)^ In addition, when considering a discharge destination from the acute setting, clinicians consider a patient’s prognosis of functional recovery as the most critical factor.^4, 5)^ Therefore, predicting a patient’s condition at discharge based on the assessment results at admission can provide a valuable contribution to personalized rehabilitation plans.

A recent review^6)^ reported that several tools had been developed to predict the outcomes for independence,^7-9)^ upper extremity function,^10, 11)^ gait,^12-17)^ and swallowing.^18, 19)^ These previous studies have predicted a single outcome based on multiple variables. However, from a clinical perspective, it is crucial to predict overall patient characteristics, incorporating multiple outcomes at discharge, not just a single outcome. Overall patient characteristics might be, for example, a patient is expected to be able to walk independently but may need assistance with communication and cognitive function. Despite the importance of predicting overall patient characteristics, previous studies have not developed such a comprehensive prediction model, leaving the problem unresolved. Therefore, developing a prediction model of overall patient characteristics at discharge in acute stroke patients will provide a new solution to a problem previous prediction models have not solved.

We employed latent class analysis (LCA) to solve these problems. LCA is a subset of structural equation modeling that categorizes heterogeneous classes (segments) from the study population by associating manifest variables with notions (latent variables) that cannot be directly measured.^20)^ LCA is useful when differences in the underlying distributions among variables contribute to heterogeneity within a study population.^21)^ Using LCA, it is possible to identify classes of overall patient characteristics (such as discharge destination, physical function, functional abilities, and length of stay) at discharge and to develop a model that predicts which class the patient belongs based on their characteristics at admission. Therefore, this study aimed to develop a prediction model of overall patient characteristics in acute stroke patients using LCA. This prediction model has the potential to predict conventional functional status and determine appropriate rehabilitation intensity levels (amount of rehabilitation per day) for individual patients.

## Methods

We conducted a retrospective observational study using the Japan Association of Rehabilitation Database.^22, 23)^ The need for informed consent was waived because all data were de-identified. The study was approved by the ethics committee of the Kanagawa University of Human Services (No. 7-20-30).

### Database

Data from the Japan Association of Rehabilitation Database, voluntarily sampled from patients admitted to participating hospitals between January 2005 and March 2016,^22-24)^ were obtained retrospectively. The data were separated into different categories based on the diagnosis and stage, such as stroke in the medical ward, stroke in the convalescent rehabilitation ward, and other conditions. The National Institutes of Health Stroke Scale (NIHSS) and Functional Independence Measure (FIM) scores, patient age, stroke type, severity, rehabilitation type, and rehabilitation length are all recorded in the stroke database. Data from 33,657 patients were collected in 2016, when 80 hospitals participated in the study. All data gathered on stroke patients admitted to acute care hospitals (37 hospitals, N=10,270) between the time of admission and discharge were used in this study.

### Participants

#### Inclusion Criteria

The inclusion criteria of this study were (1) patients with acute stroke admitted to registered hospitals in the Japan Association of Rehabilitation Database between 2005 and 2015, (2) those who were aged ≥18 years, and (3) those provided with rehabilitation (physical, occupational, and speech therapy).

#### Exclusion Criteria

The exclusion criteria were (1) no data on age at admission, (2) no data on the discharge destination, (3) age <18 years, (4) no rehabilitation provided during hospitalization, (5) duration from stroke onset to admission of >7 days, (6) rehabilitation for >9 units (180 min) a day during hospitalization, because, in the Japanese rehabilitation system, there is a limit of 9 units daily for rehabilitation. Therefore, assuming that a patient receiving >9 units a day is undergoing a normal rehabilitation program is difficult. (7) length of stay of <1 day or >179 days, (8) no data on all of the items of the FIM and NIHSS at admission and/or discharge, and (9) no data on all of the items of the FIM both at admission and discharge.

#### Outcome Variables at Discharge

Based on previous studies and clinical perspectives, we selected the following outcome variables at discharge for classification using LCA: functional abilities, cognitive functions, upper extremity function, length of hospital stay, and discharge destination. The FIM assessed functional abilities at discharge. The FIM includes 18 items to determine the amount of assistance for a person with a disability to perform daily activities safely and effectively.^25)^ All items were scored using a 7-point scale: 1, total assistance; 2, maximum assistance; 3, moderate assistance; 4, minimal assistance; 5, supervision; 6, modified independence; and 7, complete independence. Based on previous studies,^26, 27)^ we classified these seven scales into three groups: complete dependence (1–2), modified dependence (3–5), and independence (6–7). Outcome variables extracted as functional abilities were eating, transfer (bed/chair/wheelchair), toileting, locomotion (walk/wheelchair), and transfer (shower/bathtub).

Cognitive functions were also described referring to the cognitive subscale of the FIM. Three items of the FIM cognitive subscale were extracted: comprehension, expression, and social interaction. Social interaction was identified as a significant predictor of the post-rehabilitation destinations in acute stroke patients.^28)^ Patients with deficits in comprehension could constitute serious problems in their homes despite the support of their families because they might not be able to understand the provided information.^29)^

The upper extremity function was collected from the score on the motor arm, one of the NIHSS subscales. The NIHSS is a scale highly widely used in modern neurology that rates deficits.^30)^ The score of the motor arm is from 0 to 4. A high score on this item indicates severe impairment in the patient’s upper extremities.

Length of hospital stay varies from patient to patient, depending on the stroke’s severity, the patient’s background influencing their discharge destination, and other factors. Therefore, it was classified into the following four categories: 1–14 days, 15–28 days, 29–42 days, and ≥43 days.

Discharge destinations confirmed in the dataset were grouped into five groups: home, different hospitals, facilities, death, and others.

#### Predictor Variables

In this study, the predictor variables were the patient characteristics and functional ability at admission, expected to contribute to classification at discharge from previous studies and clinical perspectives. They include age, functional abilities, comprehension, upper extremity function, type of stroke, and the daily amount of rehabilitation (physical, occupational, and speech therapies) during hospitalization.

We have included the following functional outcomes in FIM items: eating,^28, 31)^ locomotion (walk/wheelchair),^32)^ and comprehension.^29, 33)^ Stroke patients had a higher possibility of being discharged home if admitted with a higher motor FIM score and a lower chance of being discharged home if they were older or had a cognitive defect.^34)^ FIM toileting was one of the initial items that predicted disposition in acute stroke patients.^31)^ Based on the motor FIM score at admission, there was a correlation between locomotion gain and other self-care in stroke patients. A variation in locomotion gain influenced the improvement in self-care.^32)^ In addition, aphasia was identified as a strong prognostic factor in post-stroke outcomes.^33)^ Furthermore, deficits in auditory comprehension, reading comprehension, and oral spelling to dictation were associated with a higher possibility of discharge to a setting other than home.^29)^

The use of the NIHSS score as an early predictor of outcome following acute hospitalization for stroke has received support.^35)^ The NIHSS score of the affected upper extremity assessed at admission was associated with long-term functional outcomes evaluated using the FIM in patients with middle cerebral artery infarction.^36)^

The types of stroke confirmed in the dataset were grouped into the following eight categories: lacunar infarction, atherothrombotic cerebral infarction, cardiogenic embolism, cerebral infarction (others/unknown), hypertensive cerebral hemorrhage, cerebral hemorrhage (others/unknown), subarachnoid hemorrhage, and others/unknown.

Regarding the amount of daily rehabilitation, following Japanese health insurance guidelines, 20-minute rehabilitation (physical, occupational, and speech therapies) is defined as 1 unit. This study categorized the amount of daily rehabilitation provided for each patient available on the dataset into four groups: <2 units (<40 min), ≥2–<4 units (40–80 min), ≥4–<6 units (80–120 min), and ≥6 units (>120 min).

#### Statistical Analyses

We used the two-step method proposed by Bakk and Kuha^37)^ in developing a prediction model of LCA for the patient characteristics at discharge. When developing a prediction model, this method has less bias than other methods and is recommended even when missing values exist.^38-40)^ The two-step method comprises the following steps: 1) to fit the latent class measurement model on its own and 2) to maximize the joint likelihood with the measurement model’s parameters and exogenous latent variables’ parameters fixed at the first step’s estimated values, allowing only the structural model’s remaining parameters to be estimated in the second step.^37)^

In the first step, we conducted an LCA with our selected outcome variables to identify the patient’s condition at discharge. The optimal latent model for the number of latent classes was determined using a combination of Bayesian information criterion (BIC), entropy, Vuong-Lo-Mendell-Rubin, and class size.^41, 42)^ For the second step, we estimated a prediction model for the calculated classes in the first step with our selected predictor variables of the patient condition at admission. In addition, we processed missing data by employing full information maximum likelihood default in Latent GOLD®6.0.^43, 44)^ We used the Latent GOLD®6.0 (Statistical Innovations, Arlington, MA, USA) for LCA and the calculation of prediction models. The significance level was set at *P*<.05.

## Results

### Patient Characteristics

After applying the inclusion and exclusion criteria in this study, 6,881 patients were included (Figure 1), and baseline characteristics at admission and discharge are summarized in Table 1. The mean age at admission was 73.7 years, and the average length of stay was 29.5 days. The total FIM scores, scores of all FIM subitems, and scores of the NIHSS improved at discharge from admission.

**Figure 1.**
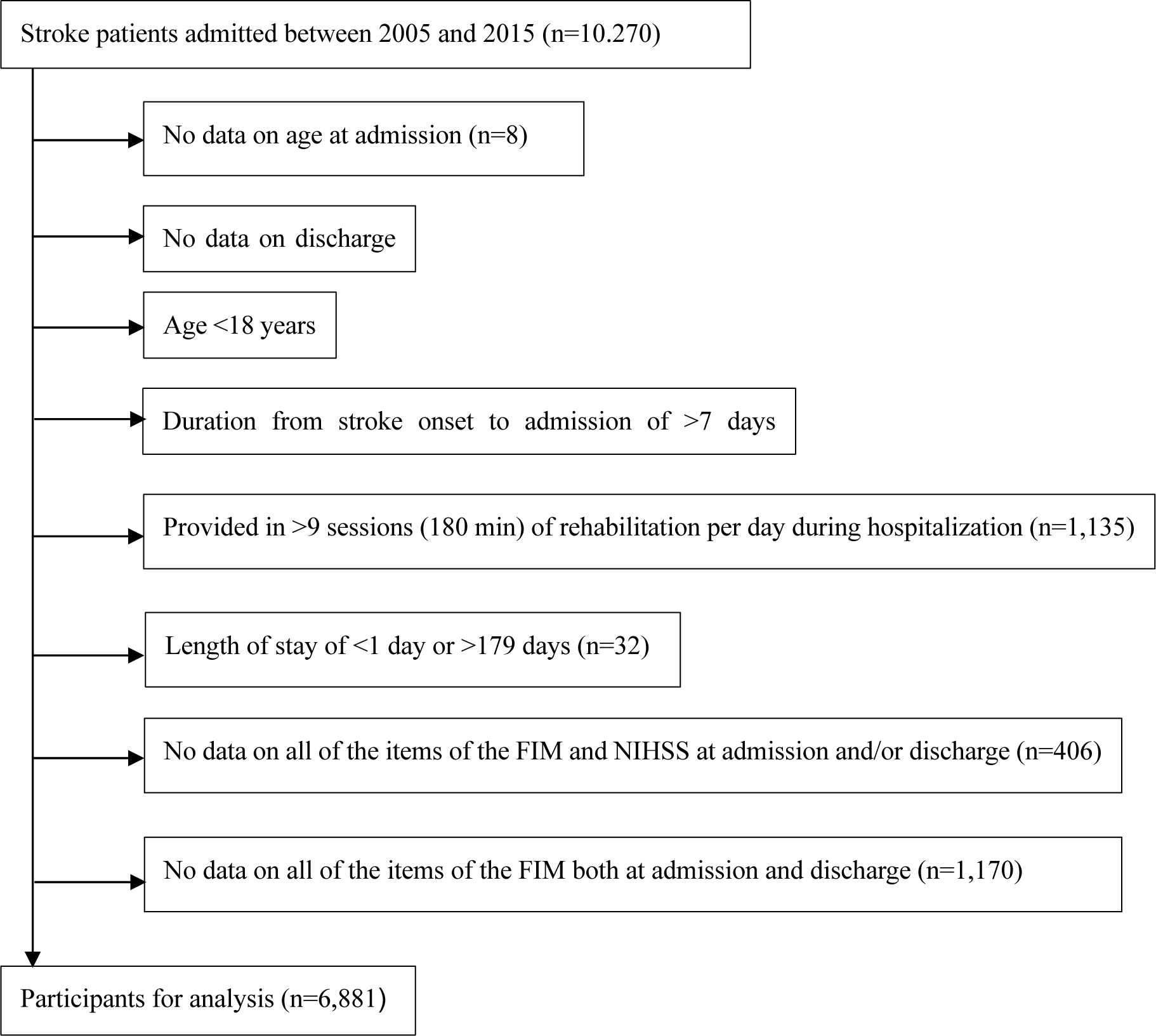
Flow diagram of the patient section process The data source was the Japan Association of Rehabilitation Database

**Table 1.**
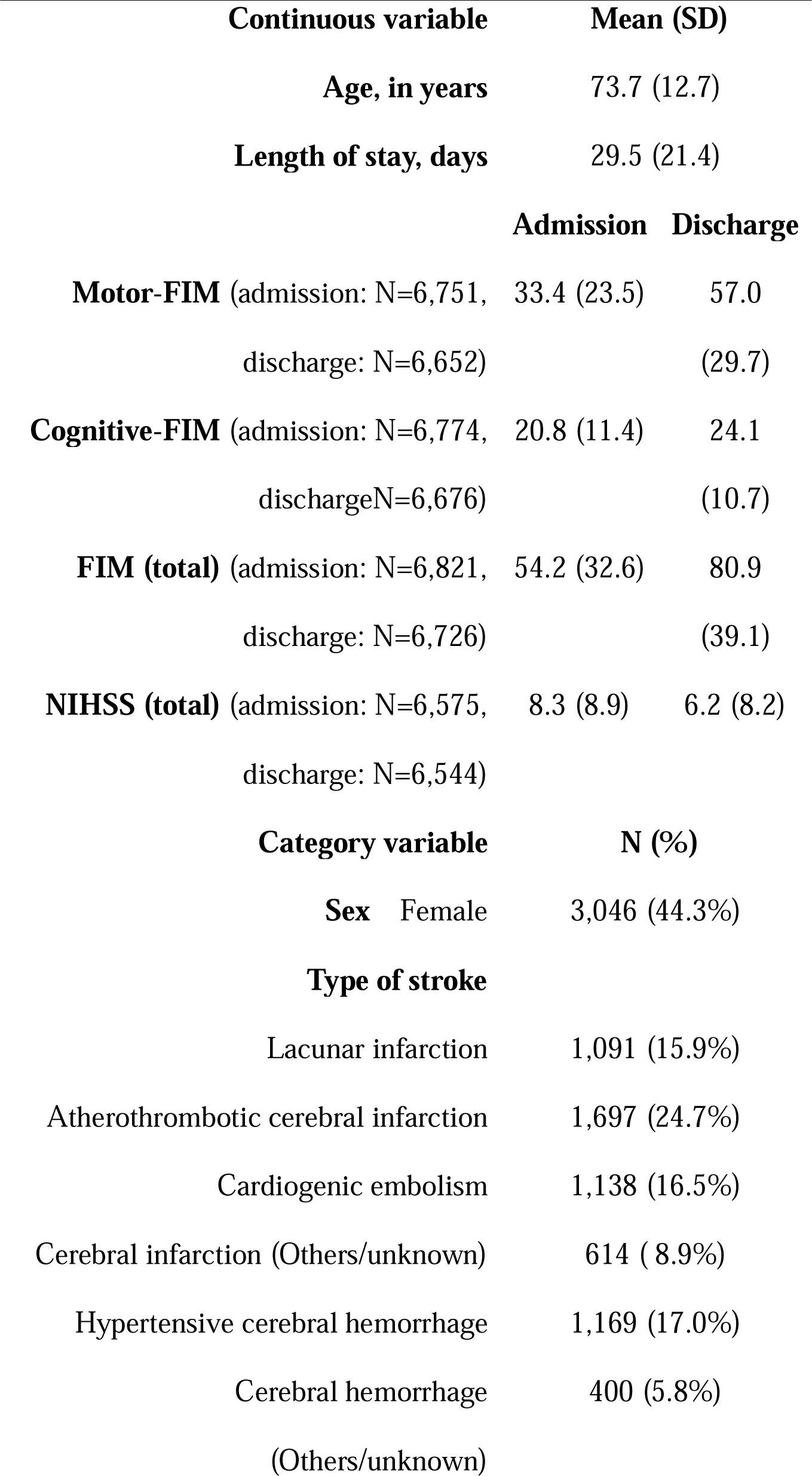
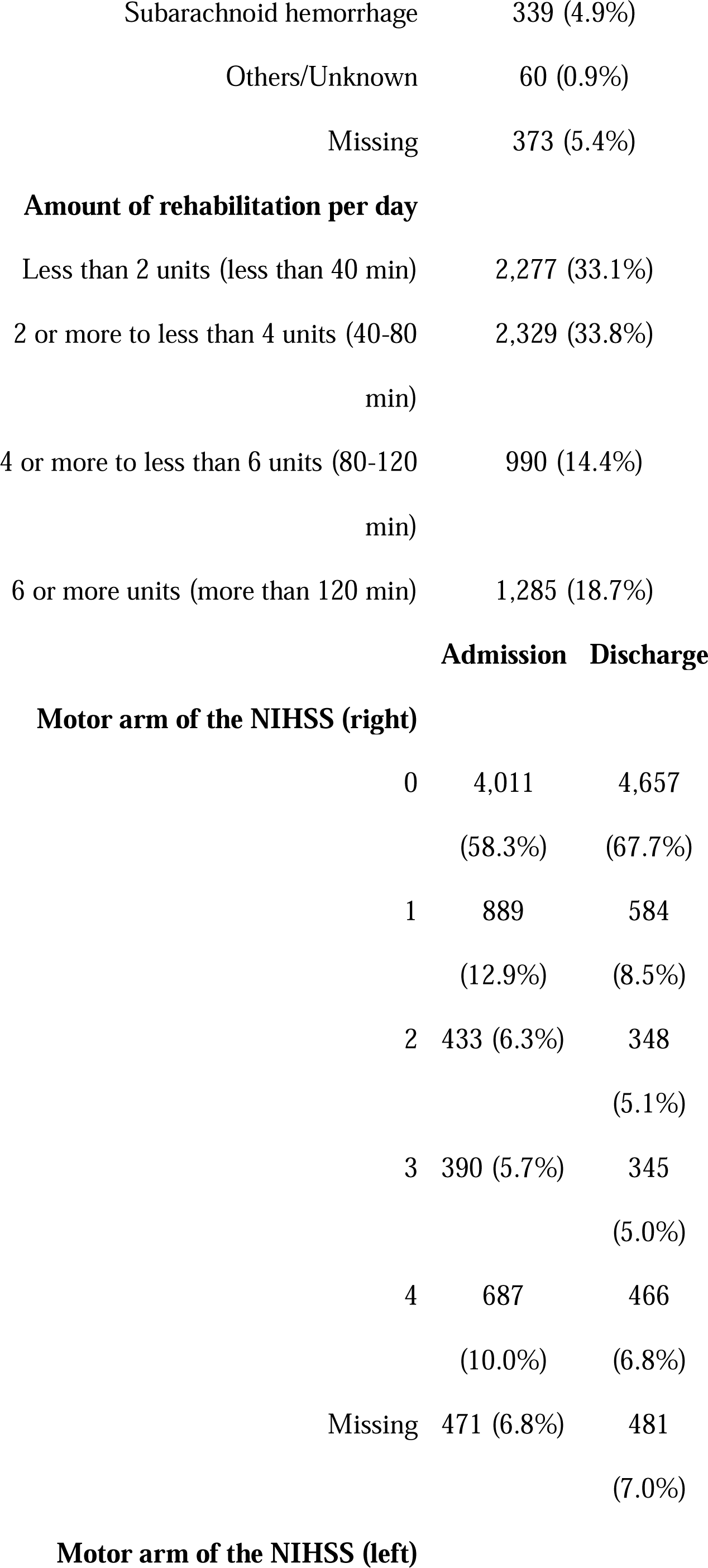
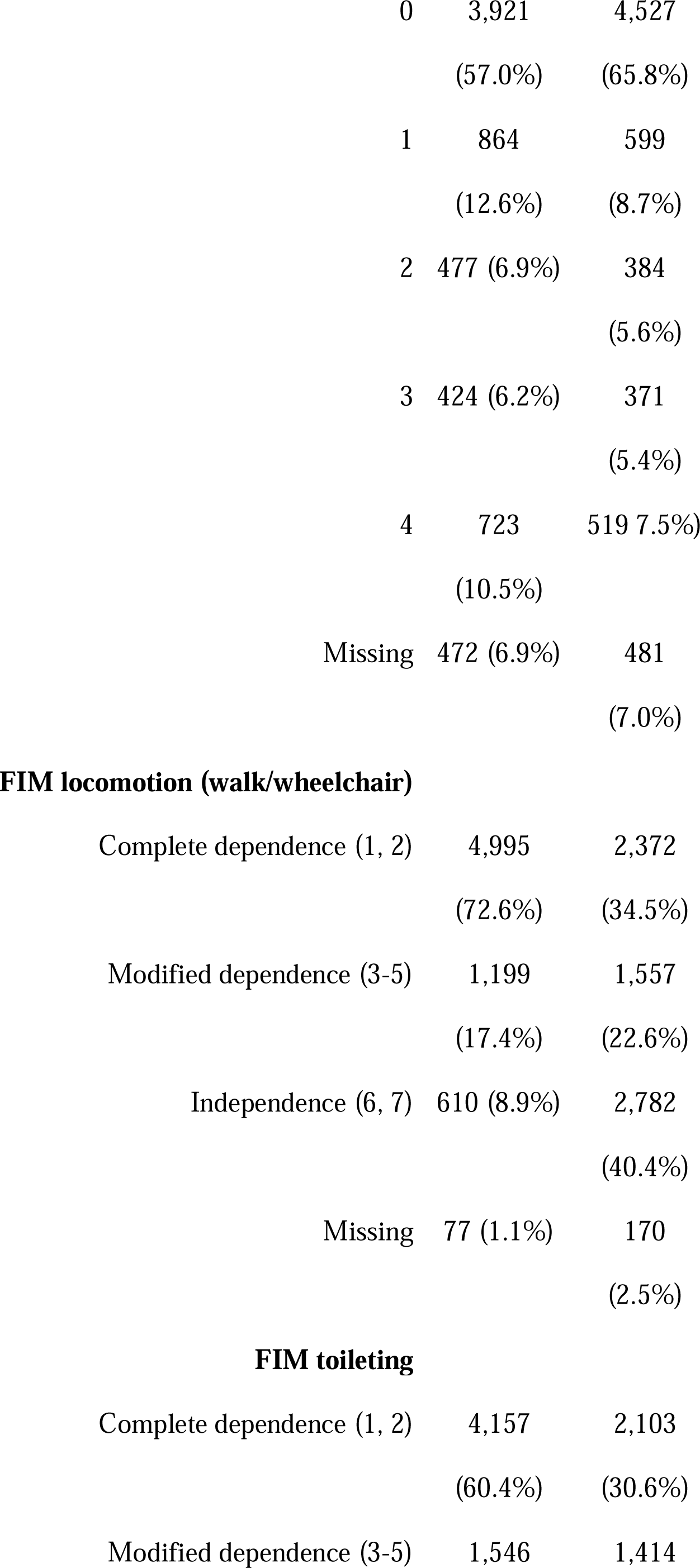
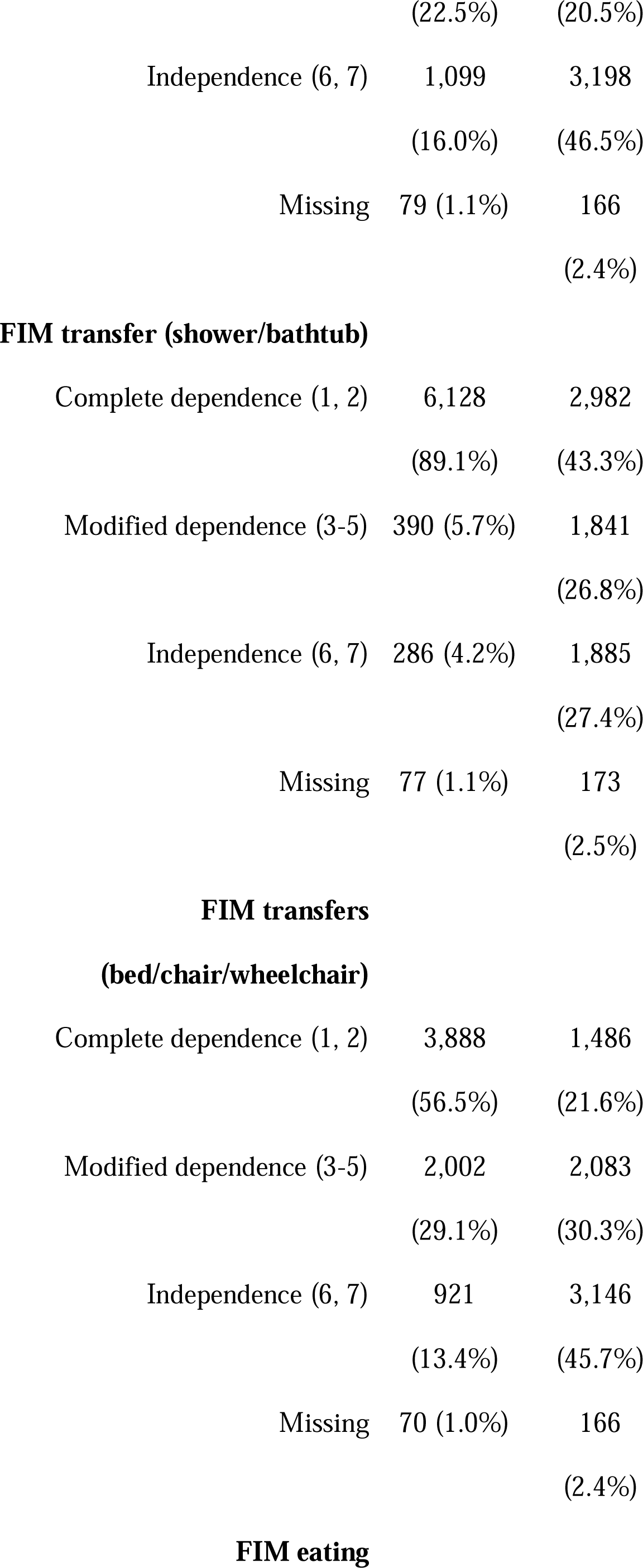
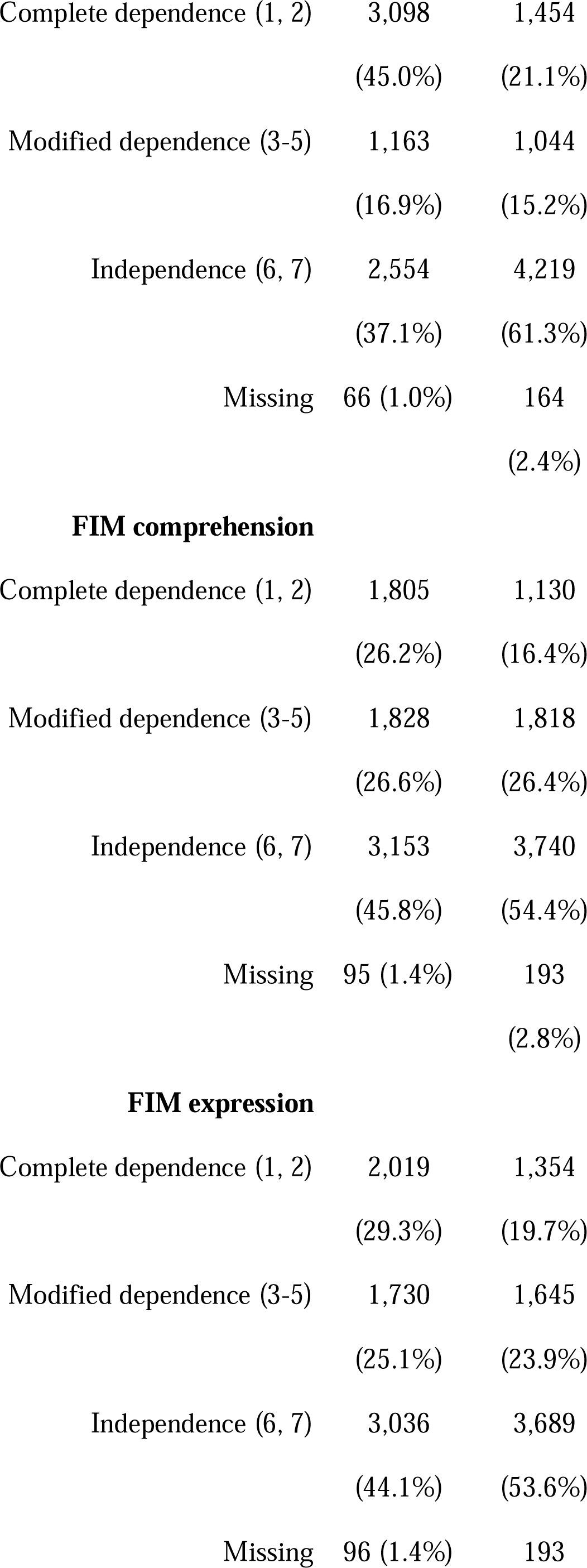
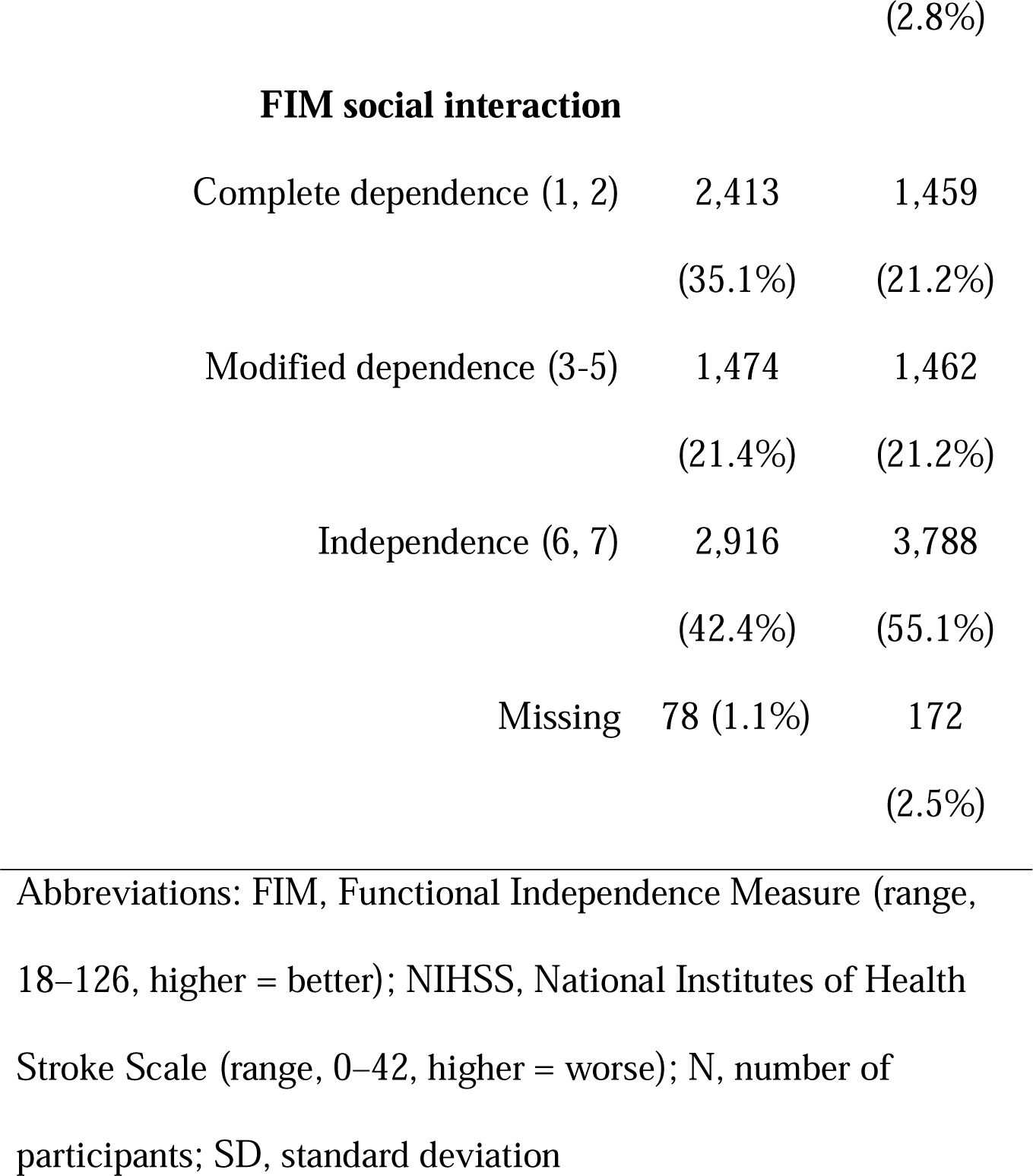
Patient Characteristics of the Study

### Latent Classes of Patient Characteristics at Discharge

Overall, 6,881 patients were classified into latent classes using LCA based on their outcomes. Latent class models with 1–12 classes were examined to select the best latent class. Model-fit statistics for LCA models with 1–12 classes are shown in Table 2. The difference in BIC/AIC fit between the models showed that it began to decline in the nine-class model. The smallest class size was <5% after the nine-class model.

**Table 2.** Model-fit Statistics for LCA Models with 1–12 Classes

| | LL | BIC | ΔBIC | AIC | ΔAIC | df | VLMR | Entropy R<sup>2</sup> | Smallest<br>class<br>size |
| --- | --- | --- | --- | --- | --- | --- | --- | --- | --- |
| 1-Class | -82978 | 166230 | - | 166018 | - | 6858 |  | 1.00 | — |
| 2-Class | -62502 | 125561 | 40669 | 125130 | 40888 | 6826 | 40952 | 0.94 | 46% |
| 3-Class | -56534 | 113907 | 11654 | 113257 | 11873 | 6794 | 11937 | 0.94 | 25% |
| 4-Class | -54963 | 111048 | 2859 | 110180 | 3078 | 6762 | 3142 | 0.92 | 16% |
| 5-Class | -53875 | 109155 | 1893 | 108068 | 2111 | 6730 | 2175 | 0.92 | 13% |
| 6-Class | -53000 | 107688 | 1467 | 106383 | 1685 | 6698 | 1749 | 0.92 | 6.8% |
| 7-Class | -52566 | 107102 | 586 | 105578 | 805 | 6666 | 869 | 0.91 | 6.0% |
| 8-Class | -52218 | 106688 | 414 | 104945 | 633 | 6634 | 697 | 0.88 | 5.6% |
| 9-Class | -51933 | 106403 | 286 | 104440 | 505 | 6602 | 569 | 0.88 | 4.5% |
| 10-Class | -51763 | 106345 | 58 | 104164 | 277 | 6570 | 341 | 0.87 | 1.5% |
| 11-Class | -51620 | 106341 | 3 | 103941 | 222 | 6538 | 286 | 0.87 | 1.5% |
| 12-Class | -51498.6 | 106380 | -39 | 103763 | 178 | 6498 | 242 | 0.86 | 1.4% |

<sup>a</sup>Based on model selection statistics, comparison of LCA models with different latent classes.
LCA, Latent class analysis; LL, Log-likelihood; BIC, Bayesian information criterion (lower values imply better model fit); AIC, Akaike information criterion (lower values imply better model fit); df, degree of freedom; VLMR, Vuong-Lo-Mendell-Rubin likelihood ratio test; Entropy, higher values imply better classification quality

In determining the number of classes, one occupational therapist and two occupational therapy students selected the nine-class model for this study, based on the interpretability of the class characteristics, given that a minimum class size of at least 5% is recommended.^42)^

Table 3 shows the class sizes and item response probabilities for the outcomes at discharge within each class. For example, a class size of 29% for Class 1 indicates that 29% of all patients belong to this class and that 97% of the patients within Class 1 would return home as “Discharge destination.” The overall characteristics of each class indicated that Class 1 was the mildest (shorter length of stay and the highest possibility of home discharge), and Class 2 was the most severe (longer length of stay and the highest possibility of transfers, including deaths). Different gradations characterized classes 3–9; these patient characteristics were clinically acceptable.

**Table 3.**
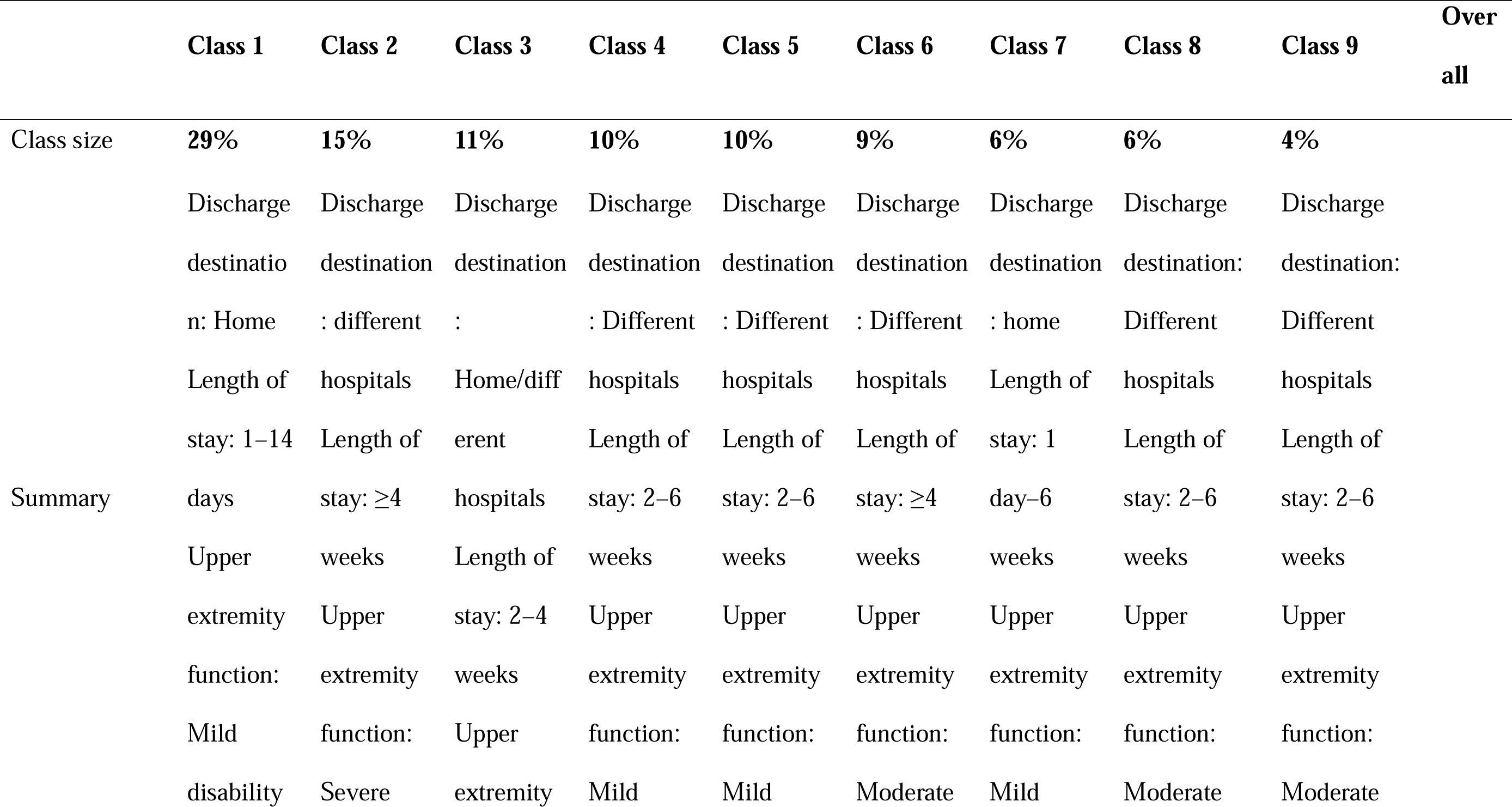
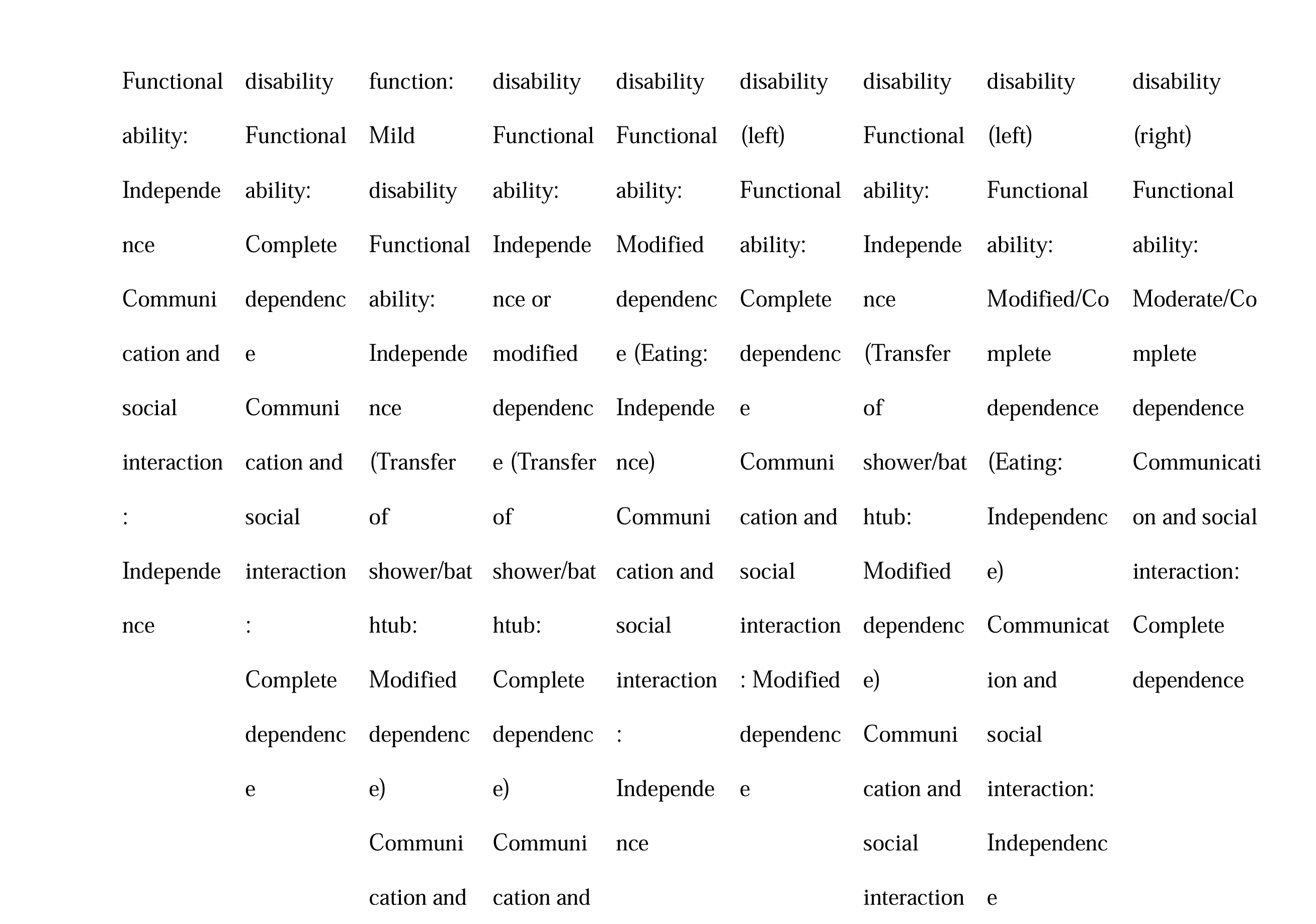
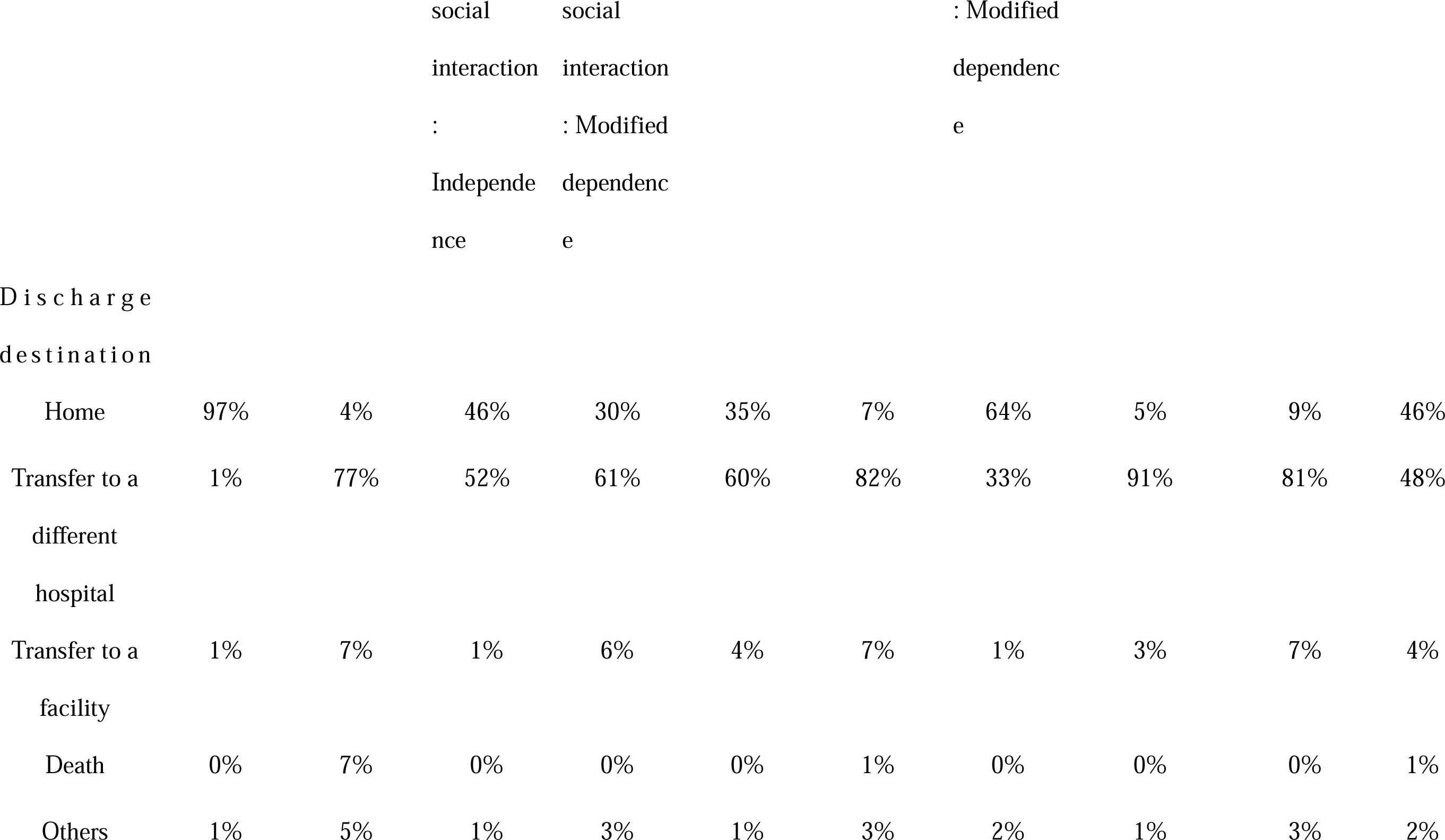
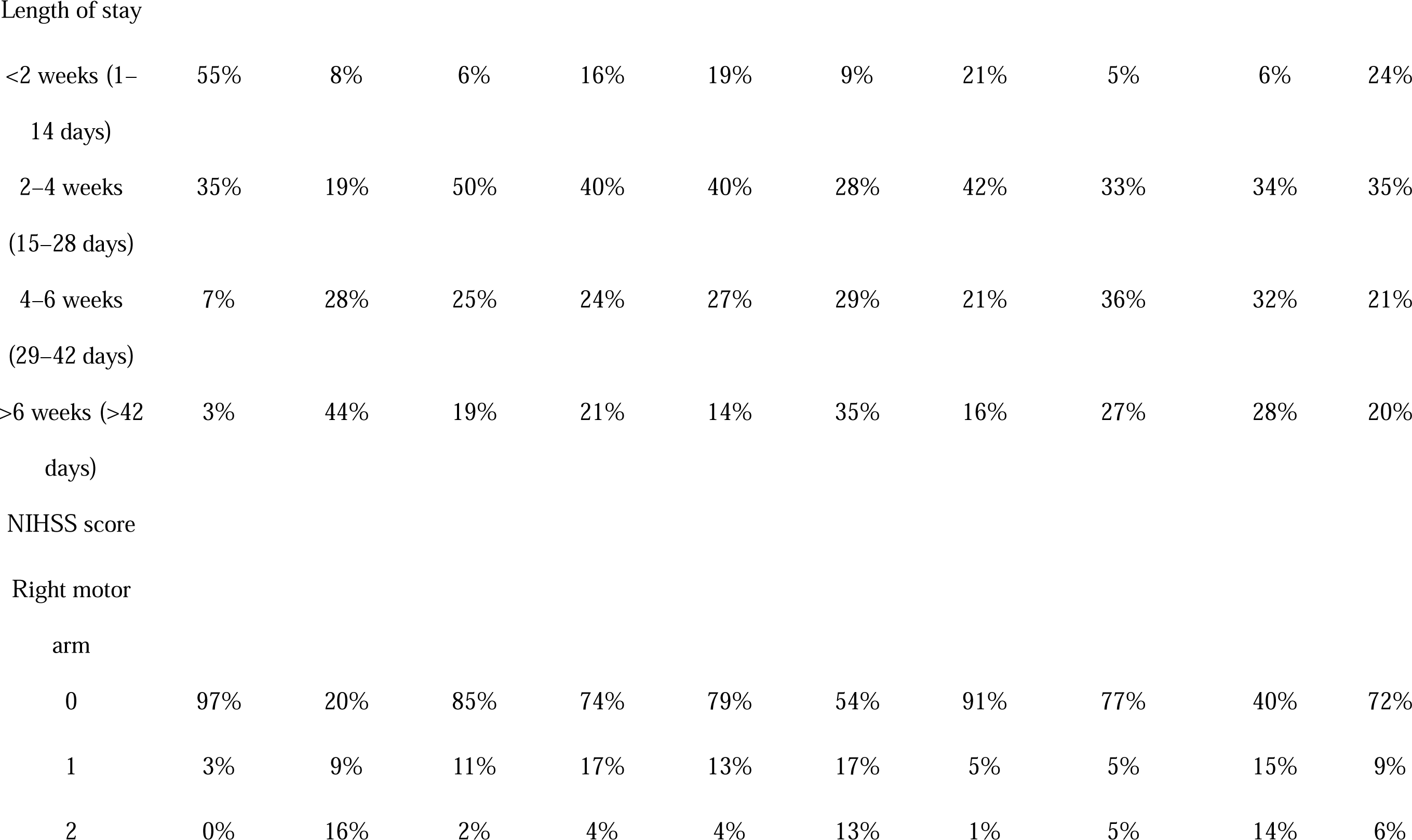
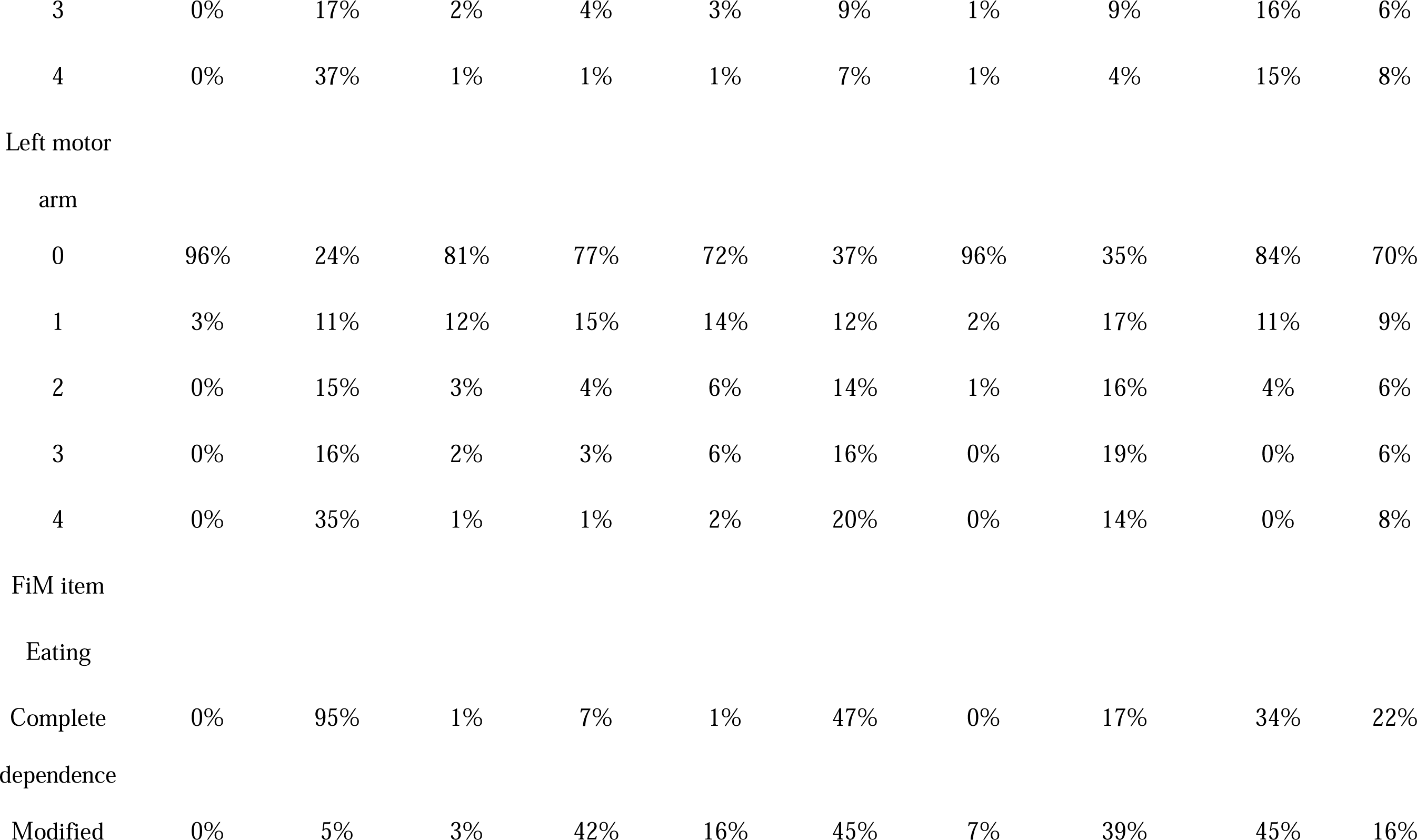
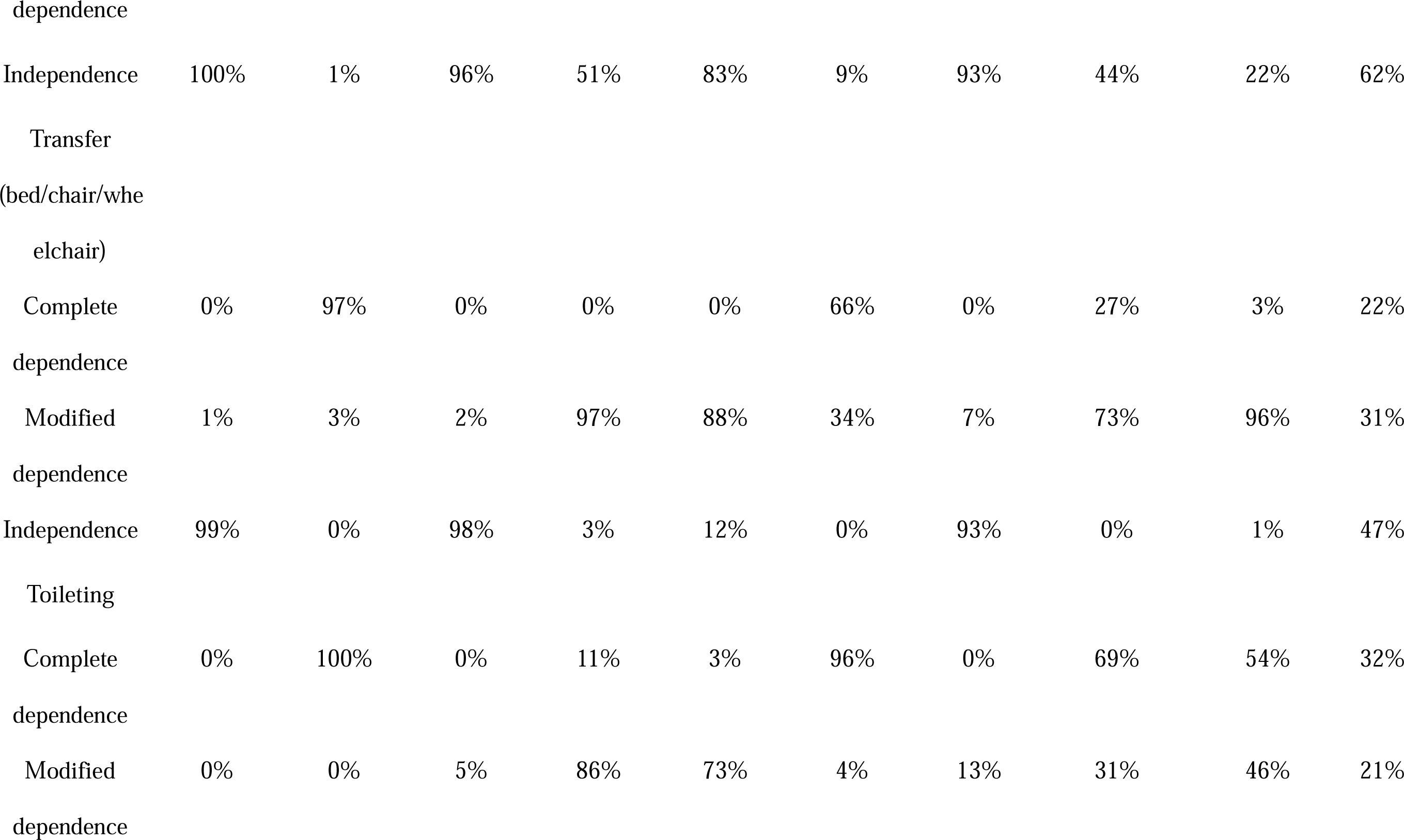
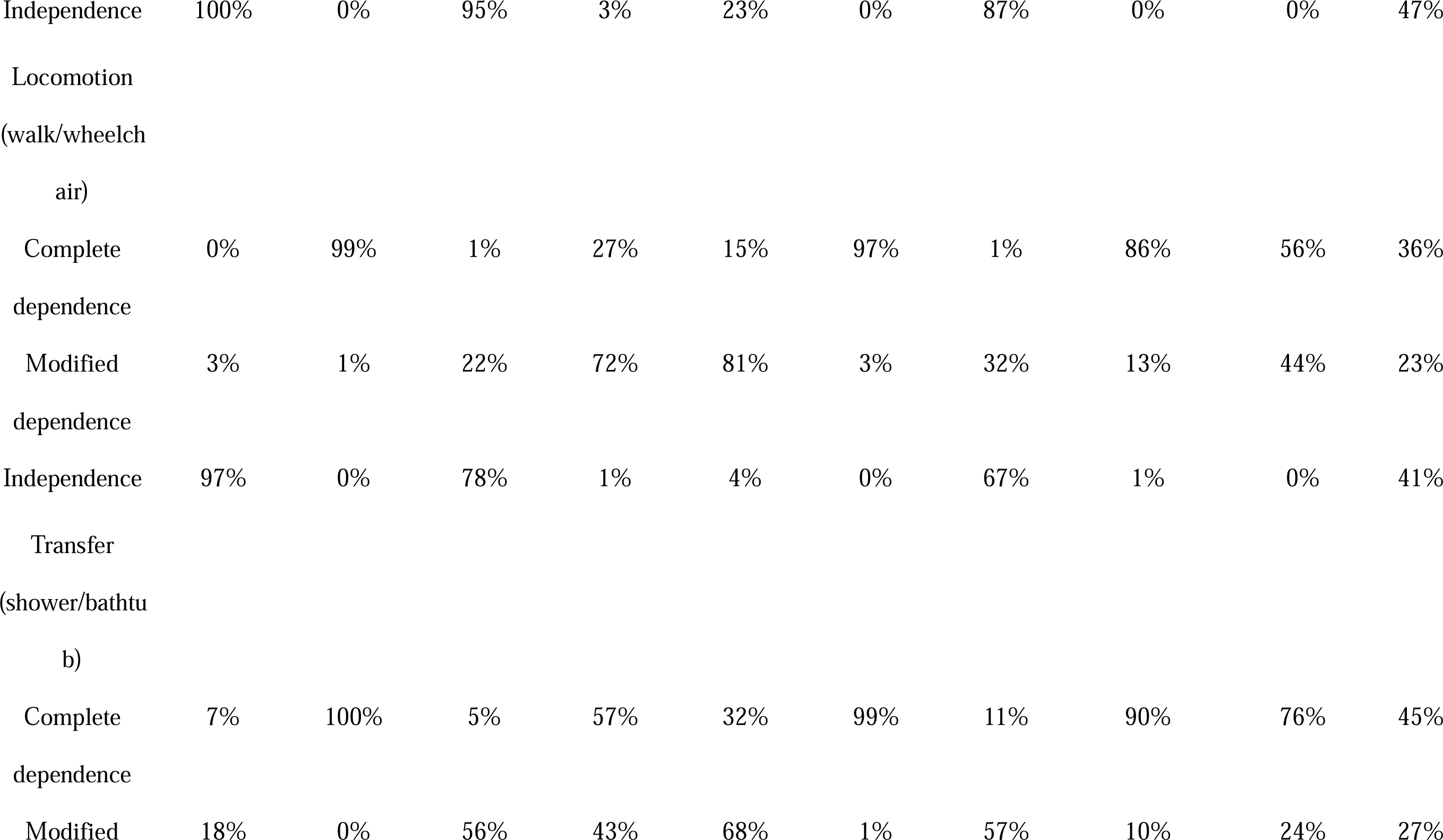
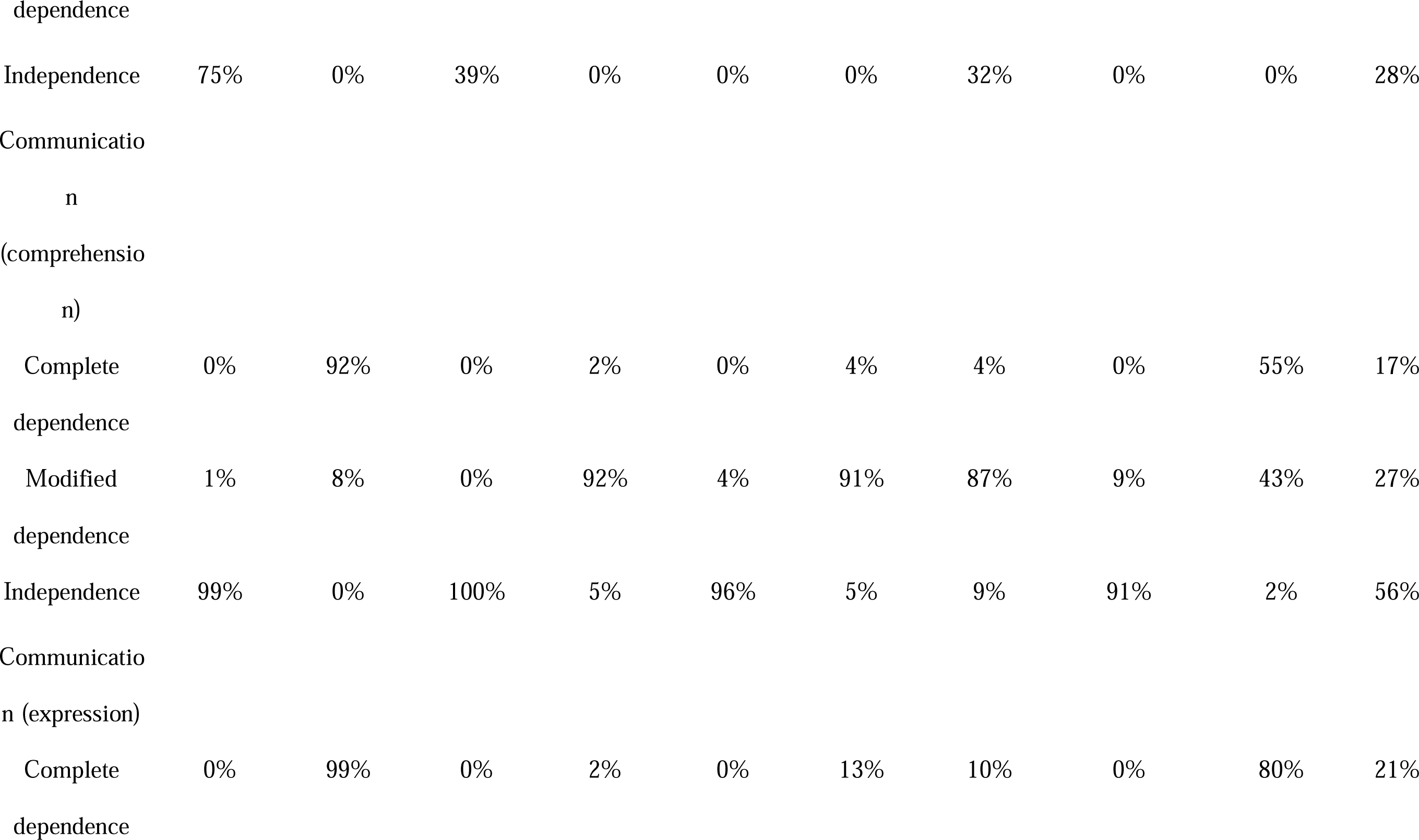
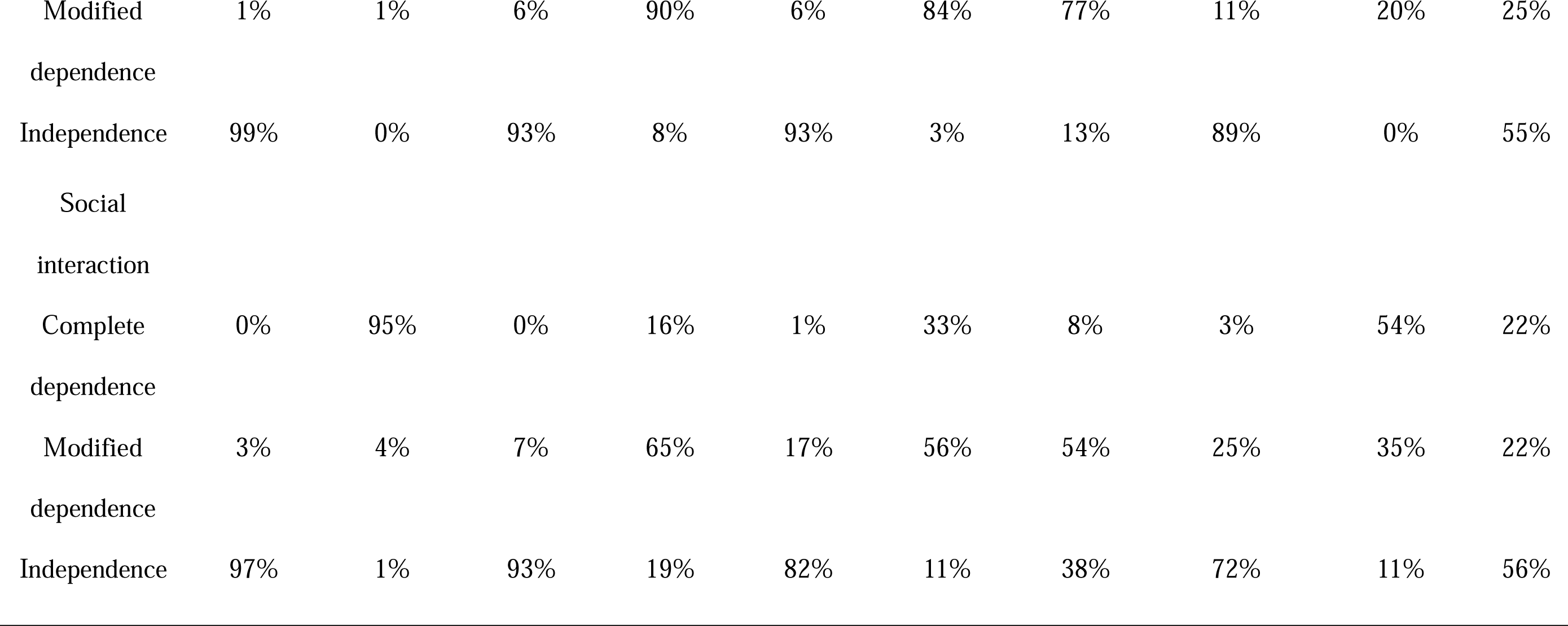
Class Sizes Belonging to Each Class and the Item Response Probabilities

### Predictors of Class Membership

All predictors of patient characteristics at admission were significant enough to predict outcomes at discharge (*P*<.01), and the standard R-squared, which indicates the goodness-of-fit of the prediction model, was 0.3383 (Table S1, S2). Relatively stronger predictors relative to Class 1 (maximum odds ratio was >10.0 in a single predictor) were the right/left motor arm of the NIHSS score. In particular, Class 8 showed extremely high odds of belonging if the left motor arm of the NIHSS score was 4. Additionally, Class 9 also showed particularly high odds if the right motor arm of the NIHSS score was 4. Furthermore, regarding the amount of daily rehabilitation, the odds ratio tended to decrease in the severe class and increase in the mild class as the amount of rehabilitation increased. However, this was not always the case.

### Model Application

Our prediction model can predict the overall patient characteristics of acute stroke patients at discharge (Video S1). The predicted results can allow practitioners to provide appropriate interventions that maximize effectiveness. Our made-up example for the model application is a patient aged 85 with a cardiogenic embolism and a severe disability on the right side of the body. The patient is provided 4–6 units (80–120 min) of rehabilitation daily and requires modified to complete assistance with ADLs. According to the predicted probability, the patient has the highest possibility of transfer and right moderate upper extremity disability at discharge. The patient is predicted to require modified to complete assistance with ADLs and complete assistance with communication and social interaction.

## Discussion

This study used LCA to classify acute stroke patients based on the overall patient condition at discharge. We also developed a model to predict their class based on patient characteristics at admission. The results showed that overall patient conditions at discharge were classified into nine classes, and all the selected predictors at admission were significantly associated with class memberships. This model was innovative in predicting the overall patient outcomes, which previous models have been unable to resolve, including the appropriate rehabilitation intensity and length of stay.

As for the predictors of this study, age, functional abilities, comprehension, upper extremity function, stroke type, and amount of rehabilitation (physical, occupational, and speech therapies) per day during hospitalization were significantly associated with class membership at discharge. These results were similar to those of previous studies.^33, 34, 36, 45)^

There are two points of particular note in this study’s findings. The first point was that the severity of the left and right upper extremities paralysis at admission had different patient characteristics at discharge. This study showed that Classes 6 and 8 were characterized as severe to moderate paralysis of the left upper extremity at discharge. Communication in these classes ranged from independence to modified dependence, although ADLs such as self-care were modified to complete dependence. These may be due to the influence of unilateral spatial neglect, which may be a factor causing the need for assistance with ADLs.^46, 47)^ In addition, Class 9 was characterized as severe to moderate paralysis of the right upper extremity at the discharge. In this class, communication was complete dependence, and ADLs such as self-care ranged from modified to complete dependence. These may be due to the influence of aphasia, which may cause difficulties with communication.^48)^ These results were clinically convincing, and appropriate rehabilitation could be provided by addressing these factors from the early stages of admission.

The second point was the intensity of rehabilitation. In some classes, a higher intensity of rehabilitation did not necessarily lead to better outcomes. It is necessary to reconsider the currently accepted philosophy “more practice is always better,” particularly in the first days after a stroke.^49)^ The model developed in this study also improves the calculation of how the overall outcome at discharge is affected by the intensity of provided rehabilitation. Therefore, by using the model of this study, providing individualized rehabilitation with its intensity optimized for the patient’s characteristics is possible.

### Limitations

There were some limitations in this study. First, we did not conduct internal and external validation. A prediction model should be validated using original data to examine internal validity through methods such as resampling (e.g., cross-validation and bootstrapping) and using other participants’ data from the same or different institutions or periods to improve the model’s validity (external validation).^50)^ Therefore, before applying this prediction model to actual clinical practice, validation of this prediction model needs to be performed. Second, the dataset we referred to did not contain enough details about interventions provided for participants. Hence, the difference in interventions among participants might have influenced the LCA and prediction model results.

Third, other potential predictors that significantly influence the outcomes at discharge can be available at admission. However, in this study, we selected predictor variables according to the findings of as many previous studies as possible and our clinical viewpoint. Therefore, we consider the model the most applicable presently. The model will be more accurate if these issues are addressed in future validation studies.

## Conclusion

In conclusion, we developed a prediction model of overall patient conditions, including the appropriate rehabilitation intensity in acute stroke patients using LCA. Before applying this model to actual clinical practice, internal and external validation would be required to improve its validity. Further studies could shed light on the potential influence of the predictors and the appropriate amount of rehabilitation for stroke patients, depending on their medical characteristics and severity.

## Supporting information

STROBE-checklist

## Data Availability

The datasets presented in this article are not readily available because of the confidential data provided by the Japan Association of Rehabilitation Database. Therefore, this data could not be shared.

## Non-standard Abbreviations and Acronyms

ADLs: activities of daily living LCA; latent class analysis
FIM: Functional Independence Measure
NIHSS: National Institutes of Health Stroke Scale

## Acknowledgments

We thank the rehabilitation patient database developed by the Japan Association of Rehabilitation Database for providing the data for this study. This study’s results and conclusions are the authors’ views, not the official views of the Japanese Association of Rehabilitation Medicine. We also appreciate the participants, their families, and the associations involved in this study.

## Funding/Support

This work was supported by JSPS KAKENHI (Grant number: JP20H03914).

## Conflict of Interest Disclosures

The authors declare
 that the research was conducted without any commercial or financial relationships that could be construed as a potential conflict of interest.

## Supplemental Material

Tables S1. Model fit of the prediction model

Table S2. Odds ratios (ORs) and p-values from the multivariate prediction of class membership (n=6,881)

Movie S1. Model application

## Notes

### Competing Interest Statement

The authors have declared no competing interest.

### Author Declarations

The study was approved by the ethics committee of the Kanagawa University of Human Services (No. 7-20-30).

### Summary of Updates

Supplemental files have been added to provide supplementary information and data supporting the findings of the study.

