## Supplemental Table 1 for "Prediction of Overall Patient Characteristics that Incorporate Multiple Outcomes in Acute Stroke: Latent Class Analysis"

**Table S1** Model fit of the prediction model

| Classification Statistics | |
|---|---|
| **Classification errors** | 0.4362 |
| **Reduction of errors (Lambda)** | 0.3924 |
| **Entropy R-squared** | 0.4167 |
| **Standard R-squared** | 0.3383 |
| **Classification log-likelihood** | -54800.7642 |
| **Entropy** | 8210.1485 |
