## Supplemental Table 2 for "Prediction of Overall Patient Characteristics that Incorporate Multiple Outcomes in Acute Stroke: Latent Class Analysis"

**Table S2.** Odds ratios (ORs) and p-values from the multivariate prediction of class membership (n=6,881)

| | Class2 OR (95%CI) | Class3 OR (95%CI) | Class4 OR (95%CI) | Class5 OR (95%CI) | Class6 OR (95%CI) | Class7 OR (95%CI) | Class8 OR (95%CI) | Class9 OR (95%CI) | p-value |
|---|---|---|---|---|---|---|---|---|---|
| **Age** | 1.2 (1.1 –1.2) | 1 (1.0 –1.0) | 1.1 (1.1 –1.1) | 1.1 (0.9 –1.1) | 1.1 (1.1 –1.1) | 1 (1.0 –1.1) | 1.1 (1.1 –1.1) | 1.1 (1.1 –1.1) | <0.001 |
| **Type of stroke** | | | | | | | | | |
| Lacunar infarction | ref | ref | ref | ref | ref | ref | ref | ref | <0.001 |
| Atherothrombotic cerebral infarction | 1.7 (0.9 –3.3) | 1.9 (1.3 –2.8) | 1.1 (0.7 –1.7) | 1.4 (0.4 –1.9) | 1.2 (0.7 –2.0) | 1.5 (0.9 –2.4) | 1.6 (0.9 –2.9) | 1.1 (0.6 –2.2) | |
| Cardiogenic embolism | 2.7 (1.4 –5.5) | 1.3 (0.8 –2.1) | 1.0 (0.6 –1.6) | 1.0 (0.4 –1.5) | 0.8 (0.5 –1.5) | 2.8 (1.7 –4.8) | 1.1 (0.6 –2.2) | 1.6 (0.8 –3.2) | |
| Cerebral infarction (others/unknown) | 3.0 (1.3 –7.0) | 1.6 (1.0 –2.6) | 1.4 (0.8 –2.5) | 1.3 (0.3 –2.0) | 1.6 (0.8 –3.2) | 1.3 (0.7 –2.6) | 1.6 (0.7 –3.3) | 1.4 (0.5 –3.4) | |
| Hypertensive cerebral hemorrhage | 2.9 (1.4 –5.9) | 2.3 (1.4 –3.6) | 1.9 (1.1 –3.2) | 1.8 (0.2 –2.8) | 2.2 (1.2 –3.9) | 1.9 (1.0 –3.4) | 2.4 (1.3 –4.5) | 1.6 (0.8 –3.4) | |
| Cerebral hemorrhage (others/unknown) | 4.3 (1.7 –10.8) | 2.9 (1.4 –5.8) | 2.8 (1.4 –5.8) | 1.9 (0.1 –3.8) | 2.1 (0.9 –4.9) | 3.1 (1.5 –6.4) | 3.0 (1.3 –6.9) | 2.1 (0.7 –5.6) | |
| Subarachnoid hemorrhage | 0.2 (0.1 –0.6) | 1.4 (0.7 –2.9) | 0.4 (0.2 –0.9) | 0.2 (0.7 –0.6) | 0.2 (0.1 –0.5) | 0.7 (0.3 –1.6) | 0.1 (0.0 –0.4) | 0.2 (0.1 –0.6) | |
| Others/Unknown. | 0.8 (0.1 –7.1) | 0.7 (0.2 –2.3) | 1.8 (0.5 –7.2) | 1.2 (0.1 –4.1) | 0.2 (0.0 –2.6) | 1.3 (0.4 –5.0) | 0.7 (0.1 –6.0) | 0.1 (0.0 –1.3) | |
| **Rigth Motor Arm of the NIHSS score** | | | | | | | | | |
| 0 | ref | ref | ref | ref | ref | ref | ref | ref | <0.001 |
| 1 | 3.2 (1.9 –5.4) | 2.3 (1.6 –3.3) | 2.6 (1.8 –3.8) | 2.1 (0.3 –2.9) | 2.8 (1.8 –4.3) | 1.6 (1.1 –2.5) | 1.7 (1.0 –2.8) | 2.1 (1.2 –3.8) | |
| 2 | 20.0 (9.4 –42.4) | 6.3 (3.3 –12.1) | 4.8 (2.3 –10.1) | 5.8 (0.0 –11.3) | 8.6 (4.1 –18.0) | 4.8 (2.3 –10.2) | 3.6 (1.4 –8.8) | 9.7 (4.1 –22.5) | |
| 3 | 42.3 (18.3 –97.7) | 4.6 (2.0 –10.8) | 3 (1.1 –8.2) | 5.3 (0.0 –11.8) | 12.4 (5.3 –29.1) | 3.9 (1.7 –8.9) | 9.5 (3.9 –23.3) | 20.4 (8.5 –48.9) | |
| 4 | 92.4 (36.9 –231.2) | 6.7 (2.6 –16.9) | 7.8 (3.0 –20.3) | 6.5 (0.0 –16.5) | 22.5 (8.9 –56.5) | 6.3 (2.5 –16.2) | 18.8 (7.2 –48.9) | 55.3 (21.8 –140.6) | |
| **Left Motor Arm of the NiIHSS score** | | | | | | | | | |
| 0 | ref | ref | ref | ref | ref | ref | ref | ref | <0.001 |
| 1 | 2.6 (1.5 –4.3) | 2.1 (1.5 –3.1) | 1.6 (1.1 –2.4) | 2.0 (0.3 –2.8) | 2.4 (1.5 –3.8) | 1.1 (0.7 –1.8) | 4.6 (2.8 –7.6) | 1.6 (0.9 –2.6) | |
| 2 | 24.1 (11.5 –50.7) | 10.6 (5.5 –20.5) | 7.2 (3.5 –14.9) | 9.7 (0.0 –18.6) | 21.1 (10.3 –43.4) | 6.6 (3.2 –13.4) | 35.1 (16.9 –72.6) | 7.6 (3.4 –17.2) | |
| 3 | 45.5 (20.5 –101.1) | 9.3 (4.4 –19.4) | 7.8 (3.5 –17.5) | 14.4 (0.0 –29.3) | 58.2 (27.5 –123.4) | 2.6 (1.1 –6.0) | 66.4 (31.9 –138.3) | 3.5 (1.2 –9.9) | |
| 4 | 61.4 (27.6 –136.8) | 7.9 (3.5 –17.5) | 6.4 (2.8 –14.6) | 9.6 (0.0 –23.1) | 64.9 (29.8 –141.0) | 2.5 (1.1 –5.7) | 107.9 (49.2 –236.7) | 1.4 (0.4 –4.7) | |
| **FIM eating** | | | | | | | | | |
| Complete dependence | ref | ref | ref | ref | ref | ref | ref | ref | <0.001 |
| Moderate dependence | 0.3 (0.1 –0.5) | 1.7 (1.0 –2.7) | 1.7 (1.1 –2.6) | 1.8 (1.2 –2.7) | 0.7 (0.4 –1.2) | 2.4 (1.4 –4.1) | 1.0 (0.6 –1.6) | 1.0 (0.5 –1.8) | |
| Independence | 0.1 (0.0 –0.2) | 0.7 (0.5 –1.1) | 0.6 (0.3 –0.9) | 0.7 (0.5 –1.1) | 0.2 (0.1 –0.3) | 1.2 (0.7 –2.1) | 0.3 (0.2 –0.4) | 0.5 (0.2 –1.0) | |
| **FIM toileting** | | | | | | | | | |
| Complete dependence | ref | ref | ref | ref | ref | ref | ref | ref | <0.001 |
| Moderate dependence | 0.4 (0.2 –0.9) | 0.6 (0.4 –0.9) | 0.5 (0.3 –0.8) | 0.6 (0.4 –0.9) | 0.1 (0.0 –0.1) | 0.9 (0.5 –1.4) | 0.1 (0.0 –0.2) | 0.2 (0.1 –0.4) | |
| Independence | 0.3 (0.1 –1.0) | 0.3 (0.2 –0.5) | 0.1 (0.0 –0.2) | 0.1 (0.1 –0.2) | 0.0 (0.0 –0.1) | 0.5 (0.3 –1.0) | 0.0 (0.0 –0.2) | 0.0 (0.0 –0.3) | |
| **FIM locomotion (wak/wheelchair)** | | | | | | | | | |
| Complete dependence | ref | ref | ref | ref | ref | ref | ref | ref | <0.001 |
| Moderate dependence | 1.4 (0.6 –3.0) | 0.6 (0.4 –0.8) | 0.6 (0.4 –1.0) | 0.4 (0.3 –0.6) | 0.2 (0.1 –0.6) | 1.2 (0.8 –1.9) | 0.1 (0.0 –0.4) | 1.0 (0.4 –2.4) | |
| Independence | 0.8 (0.2 –3.5) | 0.2 (0.1 –0.4) | 0.0 (0.0 –3.1) | 0.1 (0.1 –0.3) | 1.5 (0.6 –4.0) | 1.5 (0.7 –3.0) | 0.3 (0.0 –1.6) | 1.4 (0.1 –13.5) | |
| **FIM comprehension** | | | | | | | | | |
| Complete dependence | ref | ref | ref | ref | ref | ref | ref | ref | <0.001 |
| Moderate dependence | 0.0 (0.0 –0.1) | 1.0 (0.4 –2.3) | 0.7 (0.3 –1.3) | 1.0 (0.4 –2.1) | 0.7 (0.4 –1.5) | 0.4 (0.2 –0.7) | 1.1 (0.5 –2.5) | 0.1 (0.1 –0.2) | |
| Independence | 0.0 (0.0 –0.0) | 0.6 (0.3 –1.4) | 0.0 (0.0 –0.0) | 0.5 (0.2 –1.0) | 0.0 (0.0 –0.0) | 0.0 (0.0 –0.0) | 0.6 (0.3 –1.3) | 0.0 (0.0 –0.0) | |
| **Amount of rehabilitation per day** | | | | | | | | | |
| Less than 2 units (40 minutes) | ref | ref | ref | ref | ref | ref | ref | ref | <0.001 |
| 2 or more to less than 4 units (80 minutes) | 0.3 (0.2 –0.5) | 1.4 (1.0 –1.9) | 1.0 (0.7 –1.4) | 1.3 (0.9 –1.7) | 0.9 (0.6 –1.3) | 1.6 (1.1 –2.3) | 1.1 (0.7 –1.7) | 1.0 (0.6 –1.6) | |
| 4 or more to less than 6 units (120 minutes) | 0.2 (0.1 –0.4) | 0.7 (0.5 –1.1) | 0.6 (0.4 –1.0) | 0.6 (0.4 –0.9) | 0.3 (0.2 –0.5) | 1.3 (0.8 –2.1) | 0.6 (0.3 –0.9) | 0.7 (0.4 –1.3) | |
| 6 or more units | 0.3 (0.1 –0.4) | 2.3 (1.4 –3.6) | 2.0 (1.3 –3.2) | 1.3 (0.9 –2.0) | 0.8 (0.5 –1.3) | 1.9 (1.2 –3.2) | 1.0 (0.6 –1.7) | 2.1 (1.1 –3.8) | |

Abbreviations: FIM, Functional Independence Measure (range, 18-126, higher = better); NIHSS, National Institutes of Health Stroke Scale (range, 0-42, higher = worse); CI, Confidence interval
